# Battle with COVID-19 Under Partial to Zero Lockdowns in India

**DOI:** 10.1101/2020.07.03.20145664

**Authors:** Sakshi Babbar, Arnauv Gilotra

## Abstract

The cumulative records of COVID-19 are rapidly increasing day by day in India. The key question prevailing in minds of all is when will it get over? There have been several attempts in literature to address this question using time series, Machine learning, epidemiological and statistical models. However due to high level of uncertainty in the domain and lack of big historical data, the performance of these models suffer. In this work, we present an intuitive model that uses a combination of epidemiological model (SEIR) and mathematical curve fitting method to forecast spread of COVID-19 in India in future. By using the combination model, we get characteristics benefits of these models under limited knowledge and historical data about the novel Coronavirus. Instead of fixing parameters of the standard SEIR model before simulation, we propose to learn them from the real data set consisting of progression of Corona spread in India. The learning of model is carefully designed by understanding that available data set consist of records of cases under full, partial to zero lockdown phases in India. Hence, we make two separate predictions by our propose model. One under the situation of full lockdown in India and, other with partial to zero restrictions in India. With continued strict lockdown after May 03, 2020, our model predicted May 14, 2020 as the date of peak of Coronavirus in India. However, in current scenario of partial to zero lockdown phase in India, the peak of Coronavirus cases is predicted to be July 31, 2020. These two predictions presented in this work provide awareness among citizens of India on importance of control measures such as full, partial and zero lockdown and the spread of Corona disease infection rate. In addition to this, it is a beneficial study for the government of India to plan the things ahead.

## 1 Introduction

India reported the first confirmed case of the coronavirus infection on January 30, 2020 in the state of Kerala. The affected had a travel history from Wuhan, China. As of now, June 25, 2020, confirmed cases stand at 4.3 lakhs with more than 14,000 deaths in the country. The top 5 states with the highest number of cases include Maharashtra, Delhi, Tamil Nadu, Gujarat, and Uttar Pradesh. To protect 1.3 billion population of India from getting infection, world’s biggest lockdown in history was imposed in four phases by the government of India. The motive was to flatten the curve of the infection and slow the spread of this deadly virus. During full lockdown started from March 25, 2020, India witnessed a surge in confirmed Coronavirus cases due to Tablighi Jamaat religious congregation event held in Delhi in mid March. The meeting was estimated to have been attended by more than 5,000 members including foreigners. It was an exceptional and undesirable event occurred under the toughest restrictions executed in India. With no doubt continued full lockdown after May, 03, 2020 might have resulted in controlled spread of disease in India. But at the same time, extending it for several more weeks was not a solution either. The four phases of serious lockdown in India has brought millions below the poverty line struggling for basic needs like food and shelter. The issue of migrant workers was one of the most crucial and highlighted issue in this pandemic where millions were rendered unemployed and stranded without money, and basic needs contributing to further growth of disease in India.

It has been more than 5 months to the prevailing situation of deadly Coronavirus hounding India with rapid growth of cases. Medical researchers all over the world are busy experimenting right vaccine for the virus, academic researchers are making predictions based on machine learning and time series models and astrologers are forecasting the end of disease based on planetary position and more. The key finding everyone is trying to uncover is: when this pandemic will get over?. It is a challenging task to be addressed for the reasons that there are several time variant factors that influence spread of Coronavirus disease. This includes social distancing, population and density, public awareness, Corona testing facility, lockdown and restrictions, medical facility, nature of virus and more. Consider the factor of “social distancing” which is one way to reduce the speed of the spread of the infection through the population. Social distancing not only helps in slowing the spread of infection, but also support by keeping the peak number of cases below the capacity of the medical system. Consider Figure 1 from [13]. Where two curves of different shapes are shown. The curve on the left is a steep curve indicating exponential increase in the virus spread. With such infection rate, the local health care system gets overloaded beyond its capacity to treat people. Whereas, the curve on the right is flatter showing a slower infection rate over longer period of time. Under this situation, health care system is less stressed and the required medical attention can be given to the patients. The second curve is the output of maintaining adequate social distancing and lockdowns whereas, first curve is result of zero precautions. Hence, following social distancing and lockdowns restrictions are the key to slow the spread of the virus.

**Figure 1:**
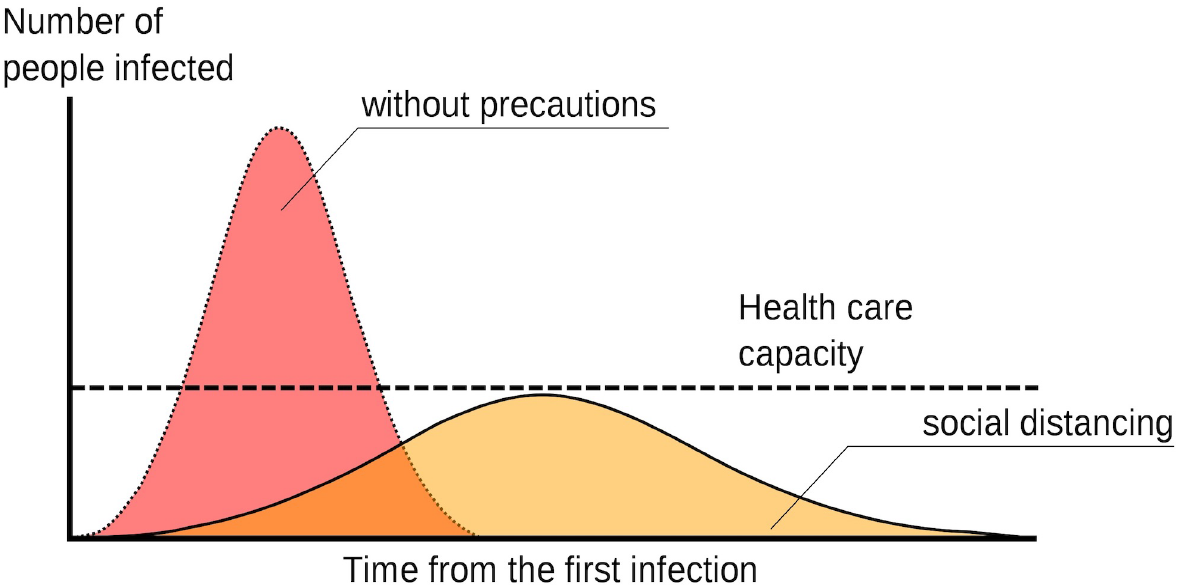
A Epidemic curve with and without social distancing

In this work, we show impact of full and partial to zero lockdowns in predicting likely end of Coronavirus in India. From full lockdown we mean the phase starting from March 25, 2020 to May 03, 2020. Where the government of India imposed strict restrictions. Whereas, partial to zero lockdown indicates the time period from May 04, 2020 to current date. Where restrictions are lighten and India is moving from locking to unlocking stage. We used a combination of epidemiological model (SEIR) and mathematical curve fitting method to forecast spread of COVID-19 in India in future. The key motivation to integrate two methods for the predictive task is to use benefits of SEIR model by making its key parameters learn using historical data of confirmed cases under full and partial to zero lockdowns in India. Fundamentally, SEIR model works by fixing the parameters such as N (population of the country/state under study), *β* (expected amount of people an infected person infects), *γ* (the proportion of infected *N* recovery), *δ*(the length of incubation period) and R_*o*_ (computed as 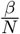 to identify lockdown scale of country/state under study) before simulation. However, we suggest that keeping these parameters fixed may affect the prediction capability of the model. Consider the parameter R_*o*_. R_*o*_ is a indicator of reproduction rate of disease. A R_*o*_ < 1 means that an outbreak is subsiding since each infected person is transmitting the virus to fewer than one other person. Whereas, R_0_ > 1 one means the virus is spreading exponentially. The scale at which virus is spreading cannot be same for every country/state. It depends upon several factors such as population and density, lockdown restrictions, rules and regulations, Corona virus testing facility and more. Hence, setting it fixed for predicting task is not a realistic solution. We propose to learn it from the data instead of keeping it fixed. On way to do is to use logistic function to learn R_*o*_. Where function is learnt from the data. Since this study is related to India so the data considered for the learning task is only meant for India. For robust learning of R_*o*_ parameter the data set is carefully used so as the right values are learnt under full and partial to zero lockdowns phases in India. The learnt R_*o*_ value is further used to evaluate parameter *β*. Once the parameters are learned, SEIR model is simulated to produce two predictions under full and partial to zero lockdown states in India. More clearly, we show end of Corona virus if the full lockdown was extended by government of India after May, 03, 2020 and, the prediction of date of peak of Coronavirus under partial to zero lockdown in India. We also show similar predictions for capital of India, New Delhi. Our propose approach has resulted in a good performance giving Mean Absolute Log Error (MALE) of 0.62 and 0.20 on fitting India’s and Delhi’s full lockdown data respectively.

The rest of the paper is organized as follows. Section 2 reviews trendy models such as time series, machine learning and deep learning for predicting Coronavirus spread in India. The summary of standard SEIR model, details on curve fitting model and our propose algorithms are presented in Section 3. The data sets used in this work are detailed in Section 4. Results achieved by our proposed algorithms are summarized in Section 5. Finally, we conclude in Section 6.

The key contributions of this work is as follows:

1. We propose a novel algorithm that is integration of standard SEIR model and mathematical curve fitting to make predictions of Coronavirus in India
2. Predict and analyze confirmed infected cases of Coronavirus in India under strict and partial to zero lockdowns.
3. To predict likely number of population to be affected in India and New Delhi by the Coronavrius.
4. Present to the citizens of India the impact of control measures such as full, partial and zero lockdowns in the spread of Corona disease infection rate

## 2 Studies in Modeling COVID-19

COVID-19 caused by deadly Corona virus is a pandemic that has spread all over the world. Under this attack by the virus, population gets infected over time. Based on immunity and other health condition, infected population can recover or die with time. The records of daily infected, recovered and deceased cases are maintained by the nations to track severity of disease. These records are further used by experts to analyze to pandemic situation and to make predictions on its possible peak of infected cases, fall of pandemic, number of deaths and recoveries over time. The key to the analysis is understanding daily trend of data making “Date” a fundamental feature to be considered to make any conclusions. This makes pandemic like COVID-19 a time series problem where trends of cases going up and down changes over time. Few examples of time series models which have been used in literature for the predicting task of COVID-19 are ARIMA, Machine learning models and Long Term Short Memory (LSTM).

In work by T. Hiteshi et. al. [1] used various time series model such as, ARIMA, single and double exponential methods and moving average to forecast future of Corona virus cases in India. The data taken for study was from January 22, 2020 to May 03, 2020. Where, observations till April 13, 2020 were used for training the models and rest were used for the evaluation purpose. It was concluded that ARIMA outperformed remaining time series models giving Mean absolute Percentage error of 4.1. It was summarized in the paper that Coronavirus cases with continue to grow in India for coming days. The study is not extended to show the prediction of ARIMA in future beyond April 03, 2020. The work is related to fitting ARIMA on the available data and showcasing closeness of model performance with actual data. Authors in [2] used Autoregressive time series models based on two-piece scale mixture normal distributions, called TP–SMN–AR models to analyze the real world time series data of confirmed and recovered COVID-19 cases. The data set comprising of confirmed and recovered Corona cases from February, 02, 2020 t0 April, 31, 2020 were considered for experimentation purpose. Model was trained on observations till April 20, 2020 and tested on remaining 10 days. The performance of model was evaluated using Mean absolute Percentage error (MAPE). The reported MAPE were 0.22% and 1.6% for confirmed and recovery Coronavirus cases respectively. Low MAPE indicated better fitting of model to the existing data. However, future predictions on development of Corona virus globally were not studied in the work. Similar work has been proposed by [3] where authors used ARIMA model on Johns Hopkins Epidemiological data to predict the CORONA epidemic trend of prevalence and incidence of COVID-19. ARIMA(1, 0, 3) and ARIMA(1, 0, 4) were discovered as best performing models to address prevalence and incidence of COVID-19 on data set used till February, 10, 2020. Authors used to these models to forecast for next two days, i.e., February 11, 2020 and February 12, 2020. The time series models ARIMA and SARIMA have also been used in study [4] on Coronavirus data set for countries Italy, Spain and Turkey. These authors also measured the performance of their model using MAPE. In addition to this, they extended their model to forecast new cases of Coronavirus in the mentioned countries. It was revealed in their work that likely decline of new cases in Italy and Spain is in July whereas; Turkey will see the decline by September 2020.

Machine learning models such as, linear regression, support vector regressor, deep neural network, recurrent neural network and Long short-term memory have also been studied to predict Coronavirus cases globally. In [5], authors used Linear regression model to forecast growth of confirmed Corona cases in India for 2 weeks from March, 30, 2020. Where linear regression model was trained on data set of the disease before March 30, 2020. The trained model was evaluated by Root mean square log error metric, which resulted in 1.75 error. It was concluded in the work that India will see growth by 5000-6000 cases during the first two weeks of April, 2020. Authors in [6] used support vector regressor, recurrent neural network and Long short-term memory for predicting task. Models were trained till observations dated April, 01, 2020 and, forecasted results were shown for next 10 days. In work proposed by A. Sina et.al. [7], machine learning and soft computing models were used to make extended prediction of 150 days using Corona virus data for countries USA, Iran and Germany. The predicted result revealed continuous progression of the outbreak in these countries. In another work by Y. Novanto [8] Long short-term memory model was used. The available global data set on Coronavirus was divided in to sets namely, training (February, 22, 2020 - March, 29, 2020) and test set (March, 30, 2020 to January, 01, 2020). Where training set was used to learn Long short-term memory model and its performance was tested on the test set. The Root mean square error was reported to be 1238.6. Authors did not use the trained model to forecast possible decline of Coronavirus cases.

Authors in [9] proposed machine learning and Cloud Computing based model to effectively to track the disease, predict growth of the epidemic and design strategies and policies to manage its spread. They fitted the available data on Coronavirus using Generalized Inverse Weibull distribution to develop a prediction framework that resulted in a statistically better performance than the baseline model (by Jianxi Luo from Singapore University of Technology and Design (SUTD)) considered in the work. Their model makes details country-wise predictions on growth and decline of Coronavirus cases. In addition to this, they have deployed their model on a cloud computing platform for more accurate and real-time prediction of the growth behavior of the epidemic. The baseline model by Jianxi Luo used in their work proposed in [10] was based on standard SEIR model. This work also presents county-wise predictions on Coronavirus cases. However, their published results on predictions do not match with current situation of Coronavirus cases globally. For example, it was forecasted by their model that COVID-19 may end in India by May, 21, 2020 but, it does not stand true. The key reason to the failure is that the uncertainty and changes are continuously evolving real-world scenarios affecting the affect the distribution of cases and hence to the curve parameters of any model.

## 3 Propose Model

The models discussed in Section 2 are intuitive enough to predict the pattern of Corona. However, they fail on revealing the likely date of maximum infected cases, the possible decline and importantly when will the pandemic ends. Probable reason to their failure is poor training of model due to lack of historical data. In addition to this, every country is different in population, diversity, density, rules and regulations and geographic structure. Data set of one country may not represent dynamics of some another country. It is not possible to have one standard training data set for a model to learn that suffice these constraints. With COVID-19 occurring globally for the first time, these time series models fails to provide desired insights. By the end of COVID-19 globally this time, each country will have enough historical data to learn and make future remedies to prevent further occurrence of this disease.

In current situation, mathematical model namely SEIR is a good choice to analyze the spread and the control of Corona infectious disease. The model categorizes each individual in the population into one of the following four groups:

1. Susceptible (S) – people who have not yet been infected and could potentially catch the infection
2. Exposed (E) - people who are exposed, but not yet infectious
3. Infectious (I) – people who are currently infected (active cases) and could potentially infect others they come in contact with
4. Recovered (R) – people who have recovered (or have died) from the disease and are thereby immune to further infections

These groups contain a certain number of people on each day. However, that number changes from day to day, as individuals move from one phase to another. For instance, individuals in compartment S will move to the compartment E, if they are exposed. Similarly, exposed people will move to the recovered I group once they catch infection. From this stage people may move to recovery (R phase) or die from the disease. The total population N across the four groups (S+ E+ I+R) is assumed to remain the same at all times. Where N is the total population of the country (or state/region) considered in the study. Since, people move from one phase to other over time, the groups S,E,I,R are functioned of time t, represented by S(t): number of people susceptible on day t, E(t): number of people exposed on day t, I(t): number of people infected on day t and R(t): number of people recovered on day t as shown in Figure 2.

**Figure 2:**
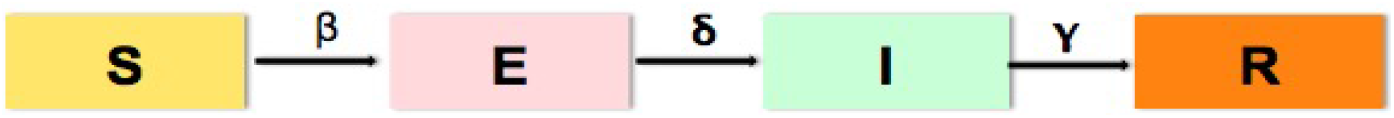
A basic SEIR Model

Based on the chain between groups, goal is to find out how the number of people in each phase changes with time. SEIR model make three simple hypotheses on what drives the movement of people between these groups. First hypothesis controls the transmission from group S to E using parameter *β*. It defines expected amount of people an infected person infects per day. Second hypothesis responsible for change from E to I is controlled by parameter *δ*. It represents the incubation period of the disease, which is the time between exposure to the virus and symptom onset. In case of Coronavirus disease it is average of 5-6 days and can be up to 14 days. Lastly, the third hypothesis, controlled by parameter *γ* is responsible is defining proportion of infected recovery per day. In summary, SEIR model consist of system of nonlinear Ordinary Differential Equations(ODE’s) in the time domain with three parameters namely, *β, δ* and *γ*. Equation 1 below represents the change in people susceptible to the disease and is moderated by the number of infected people and their contact with the infected. Equation 2 gives the people who have been exposed to the disease with time. It grows based on the contact rate and decreases based on the incubation period whereby people then become infected. The change in the infected people based on the exposed population and the incubation period is addressed by Equation 3. It decreases based on the infectious period, so the higher *γ* is, the more quickly people die/recover and move on the final stage in Equation 4.

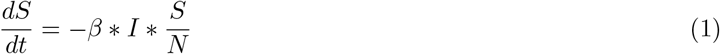

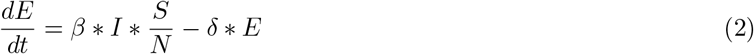

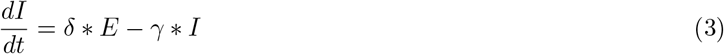

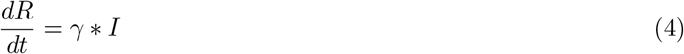

Besides three fundamental parameters in SEIR model, there is one more important variable that is important to discuss, R_*o*_ value. It is the total number of people an infected person infects. To calculate R_*o*_ value, we use the formula 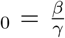. R_0_ is a indicator of reproduction rate of disease. A R_*o*_ < 1 means that an outbreak is subsiding since each infected person is transmitting the virus to fewer than one other person. Whereas, R_0_ > 1 one means the virus is spreading exponentially. R0 for Corona virus is estimated by many gropus. The Imperial College group has estimated R_*o*_ to be somewhere between 1.5 and 3.5. Most modeling simulations that project future cases are using R_0_ in that range for predictive task. GIven R_*o*_ and *γ* values from literature, *β* can be computed for Corona virus disease and Equations 1, 2, 3 and 4 are solved.

One major limitation of standard SEIR model is the user sets the parameters that control this change often based on expert knowledge and, hence remains constant during modeling. This makes the model unrealistic to capture the real trend of development of the disease. Consider the parameter R_0_ that controls the reproduction rate of the disease. Keeping this parameter constant while modeling SEIR may not be a good choice since it is influenced by several external factors. Population density, divergent demographic regions, mitigation efforts, such as social distancing, school, business, malls closures, and wearing of face masks all contribute in driving the R_*o*_ number down. In addition to this, the characteristics of Corona virus itself majorly influences on the R_*o*_ rate. In this work, we present a modified SEIR model wherein, we make the model learn SEIR specific parameters from the data instead of keeping them constant for the realistic performance. Under our proposed model, we consider actual Corona virus data related to India that details date wise confirmed, recovery and death cases. Using this data, our proposed algorithm learns parameters over time t such as: R_*o*_, *β*(t), S(t),E(t), I(T), R(t). This brings change in standard equations of SEIR model represented by Equations 1, 2, 3 and 4 as Equations 5, 6, 7 and 8 respectively.

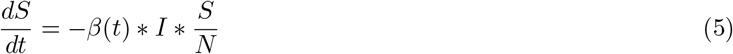

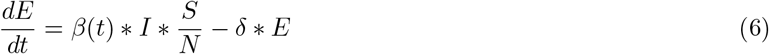

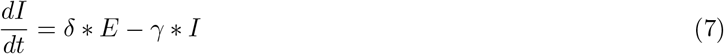

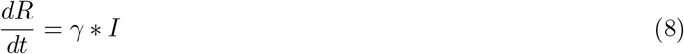

The key intuition behind learning the parameters R_*o*_ and thus *β*(t) as, 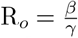 from the data is to get the real change in reproductive rate over time. As discussed earlier, the key factors influencing reproductive rate are population, lockdown controls, rule and regulations. Population of India is well known. What is unknown is how reproductive rate has grown since the first case India on January 30, 2020.

With increasing number of confirmed cases after January 30, 2020, India imposed several lockdowns to control the effect of disease. The four lockdown phases were: March, 22, 2020 - April, 14, 2020, April, 15, 2020 - May, 03, 2020, May, 04, 2020 to May, 17, 2020 and May, 18, 2020 to Mat, 31, 2020. Where first two lockdowns were very strict whereas, some relaxations were allowed in the last two lockdowns phases. The imposition of these strict and relaxing lockdowns has majorly contributed in forming the trend of confirmed Corona infected cases in India. The main idea of this paper is to automatically learn the shift in R_*o*_ value under zero, strict and relaxed lockdown phases in India and predict likely decline of COVID-19 in India. As R_*o*_ continuously changes when social distancing measures are loosened and tightened again. One choice that is adopted in this paper to capture R_*o*_ is using the Logistic function as discussed in [13]. The function is defined as in Equation 9. The summary of all those parameters that are learned and fixed in our proposed algorithm are defined in Table 1

**Table 1:**
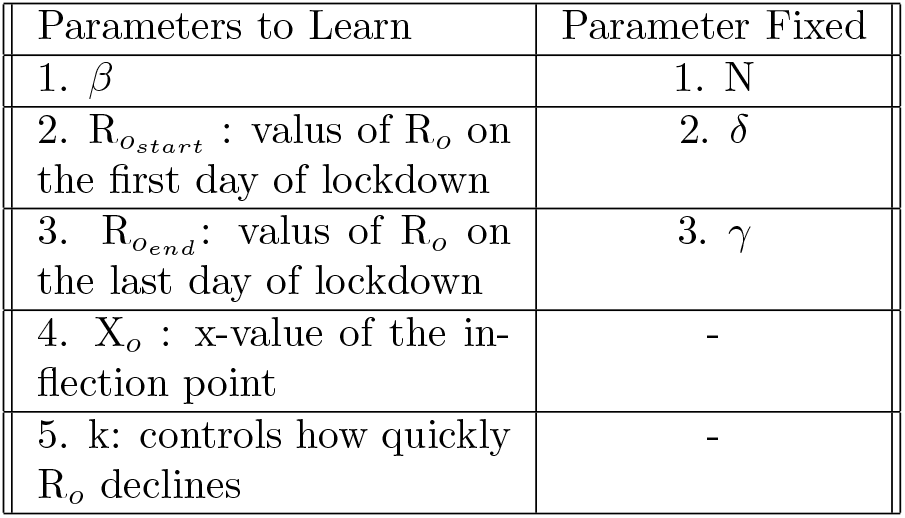
Parameters to learn Vs. Parameters fixed

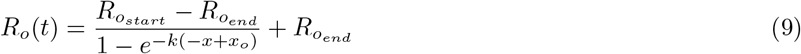

Using Equations 5, 6, 7, 8 and 9 we apply curve fitting using Coronavirus data to generate a model. Curve fitting is a process of constructing a mathematical function or a curve that best captures the series of data points given. Ideally, a good model “best fit” by capturing the underlying trend governing the data for us to make predictions of how the given data series will behave in the future. The mathematical function used in this work to learn the function f(x) from the data was “least-square”. Where the idea is to choose the parameters of the function so as to minimize the fitting error, i.e., the distance between the data values *y*_*i*_ and the y-values f(*x*_*i*_) on the fitted curve. The “least-square” method uses root-mean-square error method to compute the difference between actual and fitted value as represented by Equation 10.

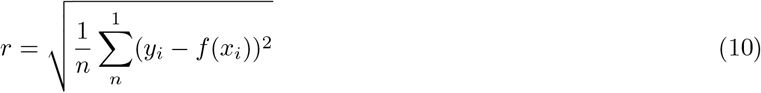

We now detail our approach of curve fitting over SEIR model using Coronavirus data in form of algorithm. For simplicity, whole process in divided into 4 algorithms. The first algorithm which we call as “SEIR_derivation” is responsible in computing derivatives of S, E, I and R over time. To compute R_*o*_ using Equation 9, we used algorithm called “*R*_*o*__derivation”. The third algorithm named “model_derivation” describes model building by providing initial parameters and computation of *β* using “*R*_*o*__derivation” function. Lastly, “Curve fitting” algorithm fits the inputted Coronavirus data using algorithms “SEIR_derivation”, “*R*_*o*__derivation” and “model_derivation”.

### Algorithm 1 SEIR_derivation

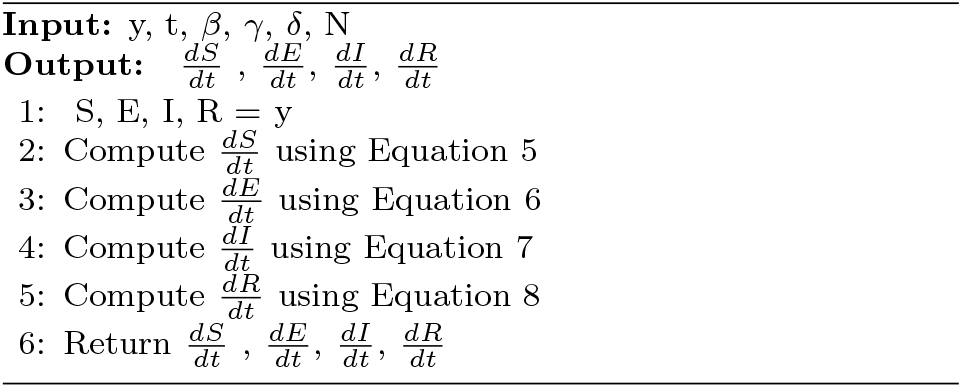

### Algorithm 2 *R*_*o*__derivation

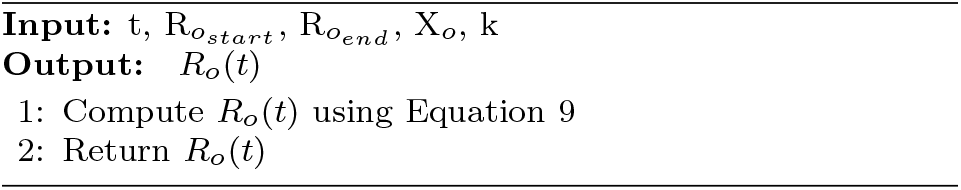

### Algorithm 3 model_derivation

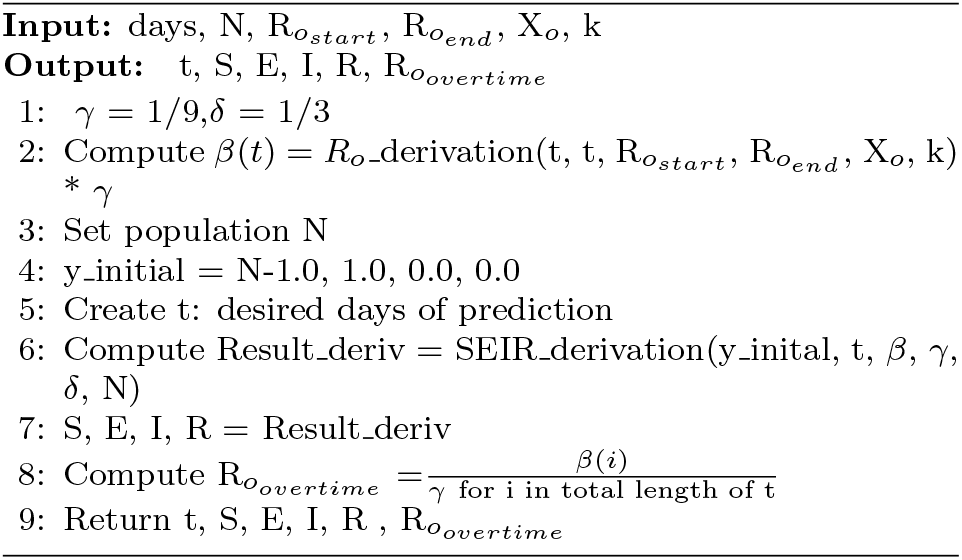

### Algorithm 4 Curve_fitting

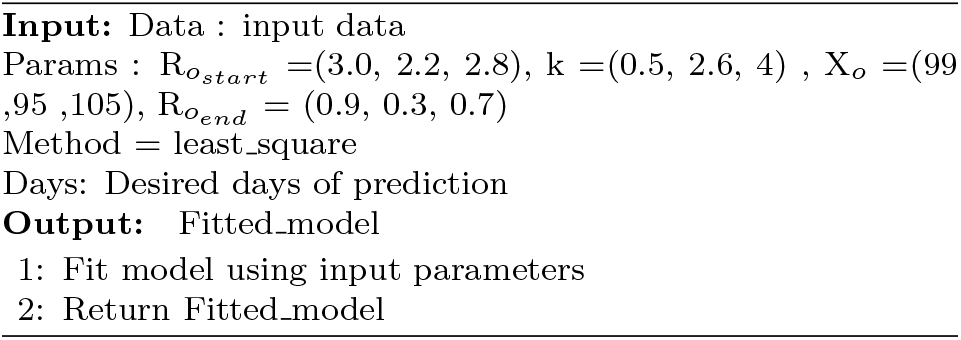

## 4 Datasets used in the Study

In this work, we used to real data set of India hosted on website [11]. As stated earlier, the aim of this work to predict confirmed Corona cases. So, we only considered data set containing date wise total confirmed, recovery, and deceased cases of individual states of India. The data set included observations from dates January, 30, 2020 to May, 03, 2020 making 96 instances. Where, January 30, 2020 marked the first case of Corona in state Kerala of India. As a first step to data preprocessing, we only kept features namely, Date, Total confirmed cases and State and deleted remaining features from the data set. The next step was to create two data sets from this main data set representing exclusive total confirmed cases of India and Delhi respectively. This step was required since the aim was to develop predictive modeling for India and Delhi separately on COVID-19 confirmed cases. In this present study, we considered India and Delhi to showcase predictive results. However, our propose model is robust enough to be deployed on data set on other states for the predictive task.

The above preprocessing steps resulted in two main data sets. One data set containing date wise cumulative confirmed cases in India starting from January 30, 2020 to May, 03, 2020. Second data set represents Delhi from February 02, 2020 to May 03, 2020 with 64 records. Figures representes tend of daily-confirmed cases in India and Delhi for 96 and 65 days respectively.

For each data set, we performed two main predictive analyses considering strict and partial to zero lockdown. During the time period from March 22, 2020 to May, 03, 2020, India was under strict lockdown phase. However, after May 03, 2020, government of India reduced the strictness and eventually it became situation of zero lockdown. This gap between strict lockdown to almost zero lockdown has given rise to several fluctuations in CORONA confirmed cases. Considering these fluctuations and to provide model the true behavior of confirmed cases under strict and zero lockdowns, we divided each data set in two parts. The period from till May, 03, 2020 in each data set was considered as strict lockdown whereas, remaining data from May, 03, 2020 was considered to be relaxing period. Under strict predictive analysis, we made the model learn the trend of confirmed till May 03, 2020 and forecasted its performance for the next 180 days. This analysis revealed the peak of confirmed Corona cases conditioned on continued strict lockdown by the government. In case of relaxed lockdown, we made the model learn from the remaining time period left in the data set and discovered the peak of Corona cases.

## 5 Results

In this section, we present experimental results on predicting peak of Coronavirus cases in India and Delhi under two main situations: (a) under strict lockdown and, (b) partial to zero lockdown. Under strict lockdown phase, observations till May 03, 2020 were considered for all different data sets used in this work for model learning. And, predictions were made for next 180 days using the learned model. The outcome of this experiment revealed situation of virus spread in India under continued strict lockdown beyond May 03, 2020. Besides extended predictions of 180 days under strict lockdown situation, we also present the outcome of model fitting on strict lockdown data set using algorithms discussed in Section 3.

Figures 5 and 6 represents fitting of learned model over actual new cases of Coronavirus data set of India and Delhi respectively. The curve fitting outcome on data set of India in Figure 5 shows similar trend of actual increase in new cases of Coronavirus infection till May 03, 2020. We notice, steep increase in cases from start of April in actual new cases (as indicated by blue line) whereas, the predicted is increasing after mid of April. The one major reason of this gap is because of religious congregation “Tablighi Jamaat” happened during strict lockdown in India. Under strict lockdown such events are not expected to occur. The actual number of new cases till May 03, 2020 in India were 40124 whereas, our model it 33463. Mean squared log error (MSLE) computed between actual and predicted cases was 0.62. Low MSLE indicates that our algorithm has fitted the actual trend of cases very well and, can be used to forecast for upcoming days. Similarly, the model fitting on data set of Delhi reveals that our predicted results are very close to actual data of new cases of virus infection. Figure 6 shows outcome on data set of Delhi. The actual cases till May 03, 2020 in Delhi were 4122 whereas, model predicted it be 3599. The MSLE for this data set was computed to be 0.20.

**Figure 3:**
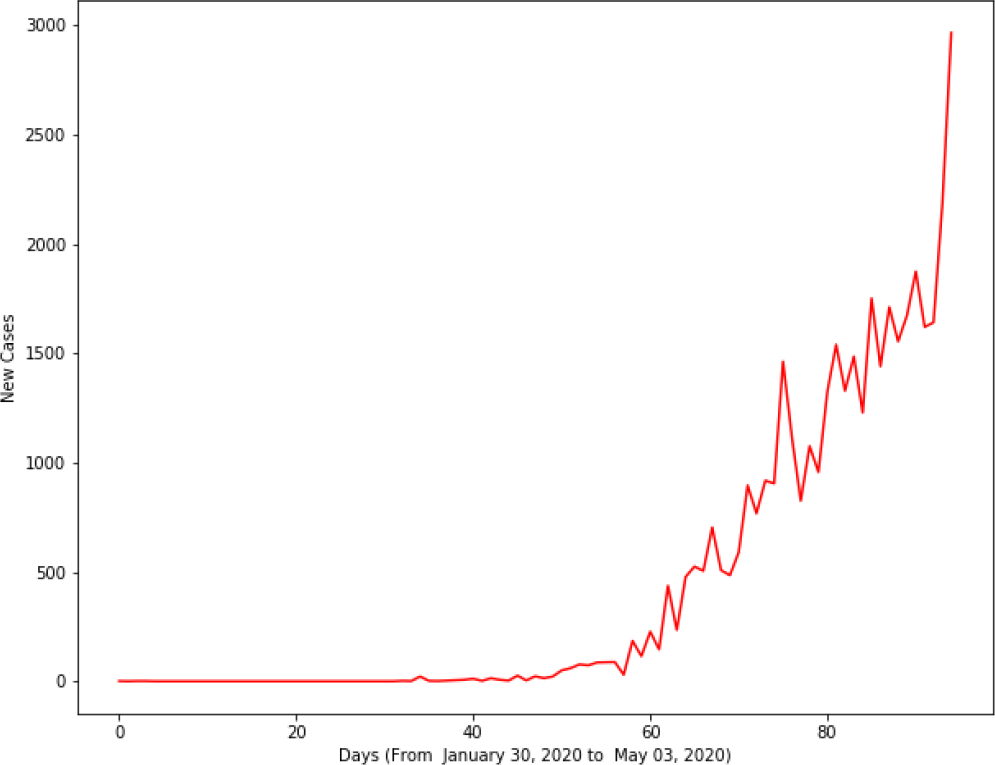
Daily new cases in India from January, 30, 2020 to May, 03, 2020

**Figure 4:**
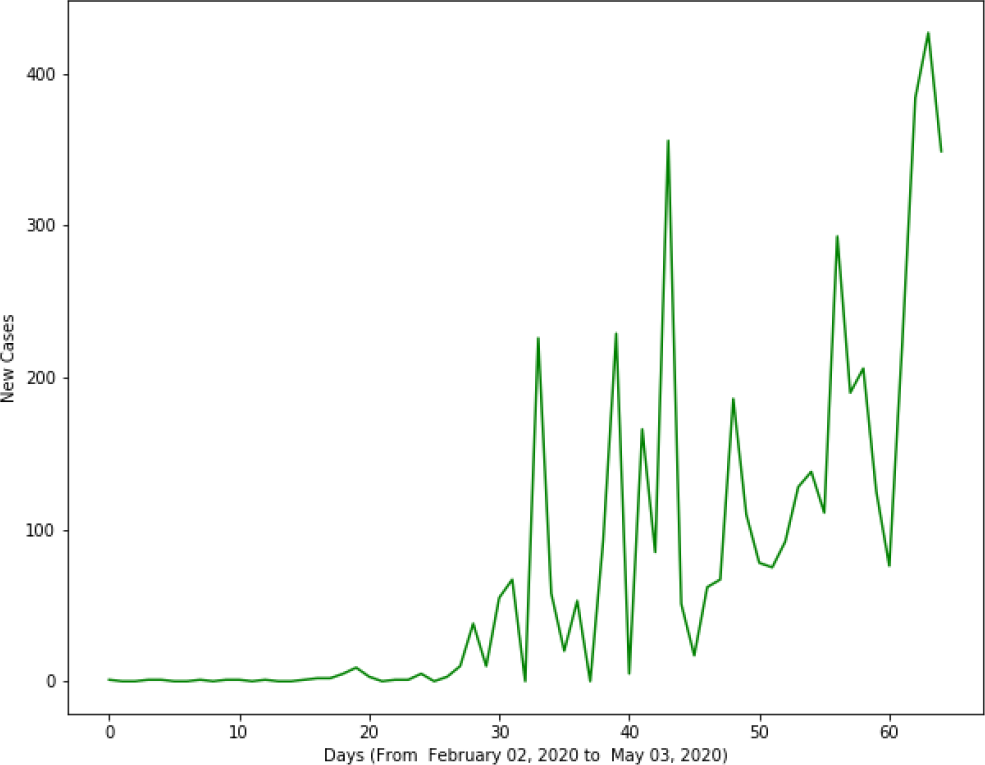
Daily new cases in Delhi from Feburary, 02, 2020 to May, 03, 2020

**Figure 5:**
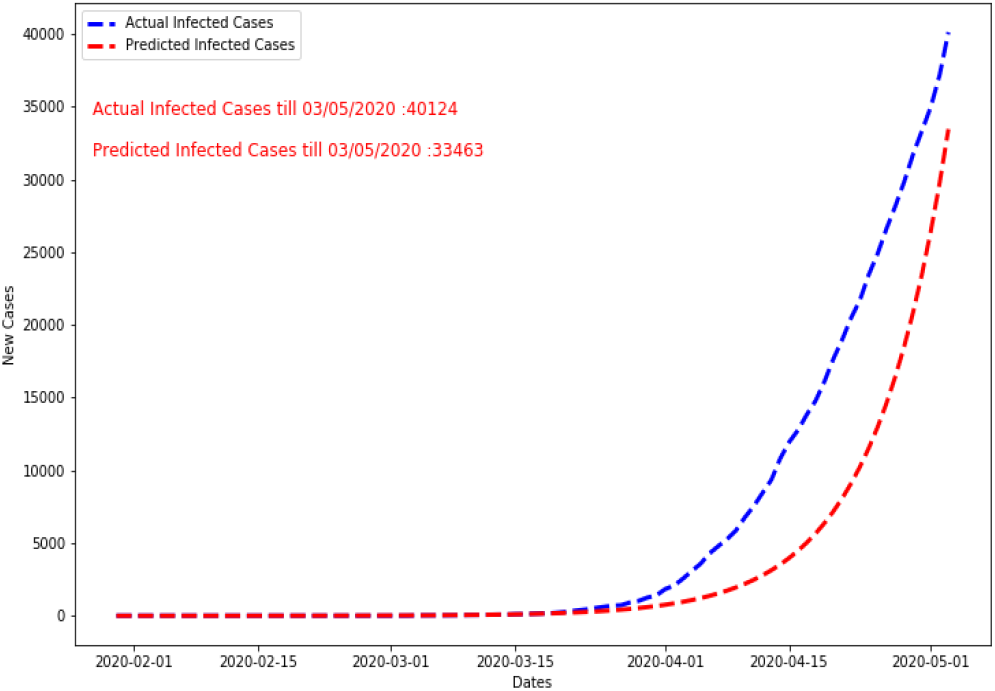
Actual Vs. Predicted New cases In India under Strict lockdown (March 22, 2020 to May 03, 2020)

**Figure 6:**
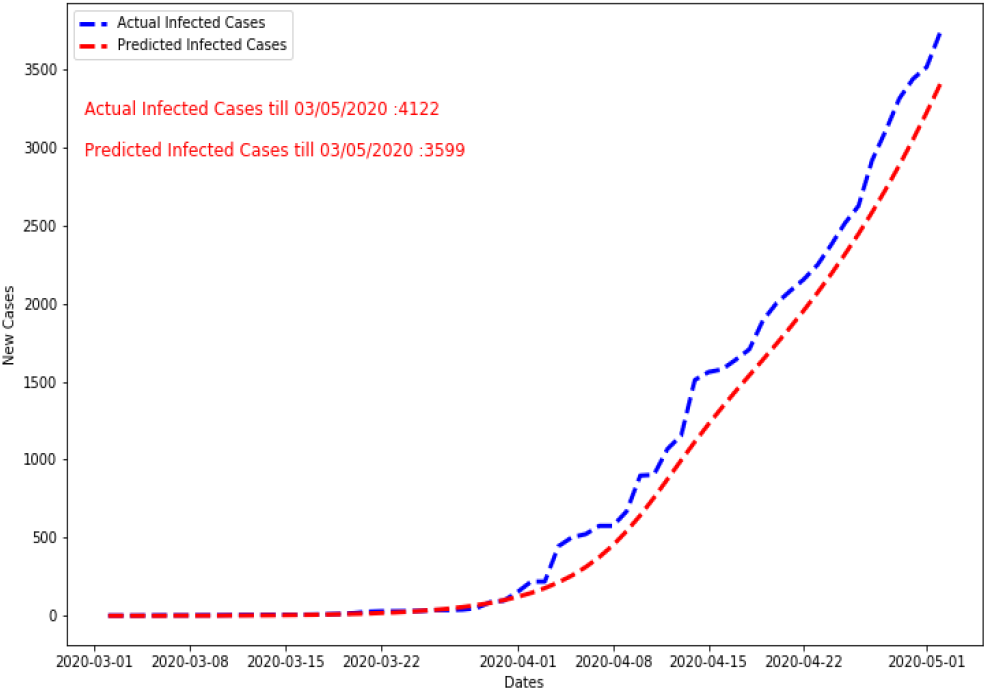
Actual Vs. Predicted New cases In Delhi under Strict lockdown (March 22, 2020 to May 03, 2020)

We now present predictions on trend of new cases of Coronavirus for continued strict lockdown beyond May 03, 2020 to discover its peak and decline. The fitting of real data set on our propose algorithm revealed that predicted peak in India was on May 14, 2020and thereby was decline in new cases. In Delhi peak predicted was May, 08, 2020. Figure 7 and Figure 8 shows the peak and decline trend of new cases in India and Delhi respectively. The 95% of Corona virus effect was expected to vanish by early, July 2020 conditioned on continued strict lockdown. Table 2 summarizes values of parameters 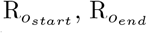, k and X_*o*_ learnt by the model on data sets of India, Delhi and Haryana. As per the results, peak of new cases were predicted on 91^*th*^ day from start of spread of disease in India and 41^*th*^ day in Delhi.

**Table 2:**
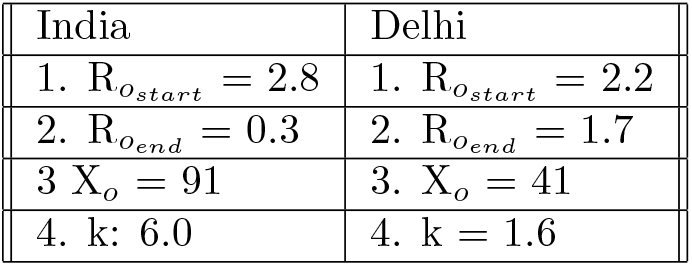
Learnt parameters over data set of India and Delhi under strict lockdown

**Figure 7:**
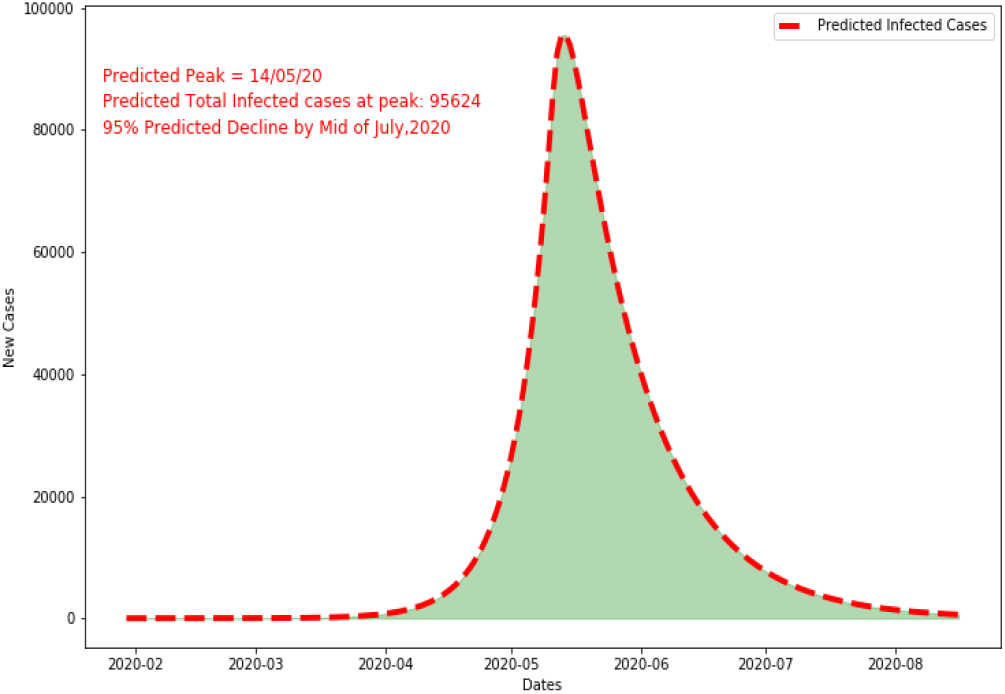
Predicted Peak of New cases In India under Strict lockdown (March 22, 2020 to May 03, 2020)

**Figure 8:**
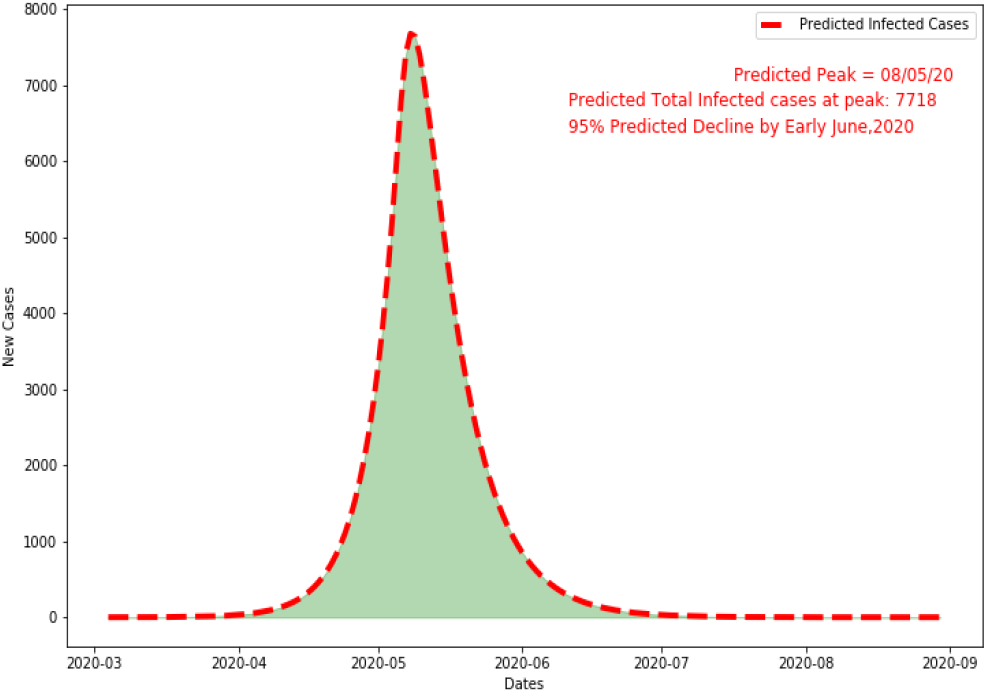
Predicted Peak of New cases In Delhi under Strict lockdown (March 22, 2020 to May 03, 2020)

The above-presented results were to stand true if there was a situation of extended strict lockdown after May 03, 2020 in India. However, it is well known that relaxation in lockdown have been eased steadily after May 03, 2020 and, has reached to partial to zero lockdown stage in India. Considering this, data available till today may not be a good choice for a model to learn since it is the combination Corona cases occurred during of strict and zero lockdown phases in India. Hence, to predict the situation of new cases of Coronavirus in India, it was important to discard the records before May 03, 2020. The key assumption made by the model to make new forecast for India was to consider relaxed lockdown. We made predictions for 300 days by our proposed model with values of key parameters 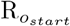 and 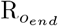 set manually instead of learning them from data. The results of prediction are shown in Figure 9 and Figure 10 for India and Delhi respectively. As per the predictions by the model, the peak of Coronavirus is expected to hit the nation on July 31, 2020 with over 1562351 cases and thereby declining. The 95% cases are forecasted to vanish by September, 2020. In Delhi the disease will be at peak on July 27, 2020. The model was fiited with 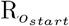 of 5.0, for India and 4.5 for Delhi.

**Figure 9:**
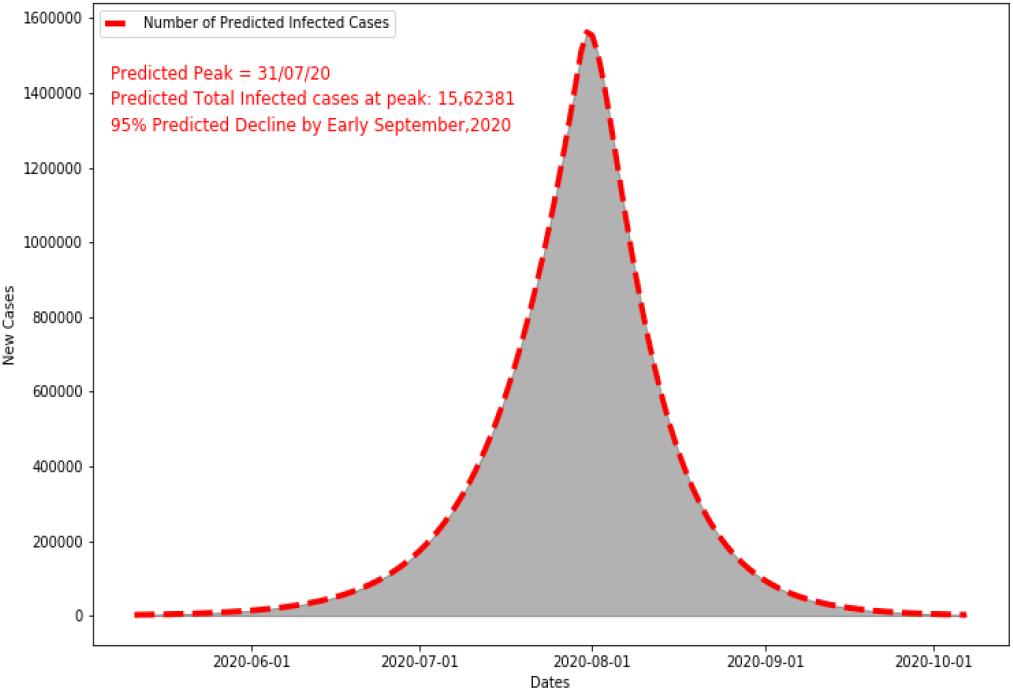
Predicted Peak of New cases In India under

**Figure 10:**
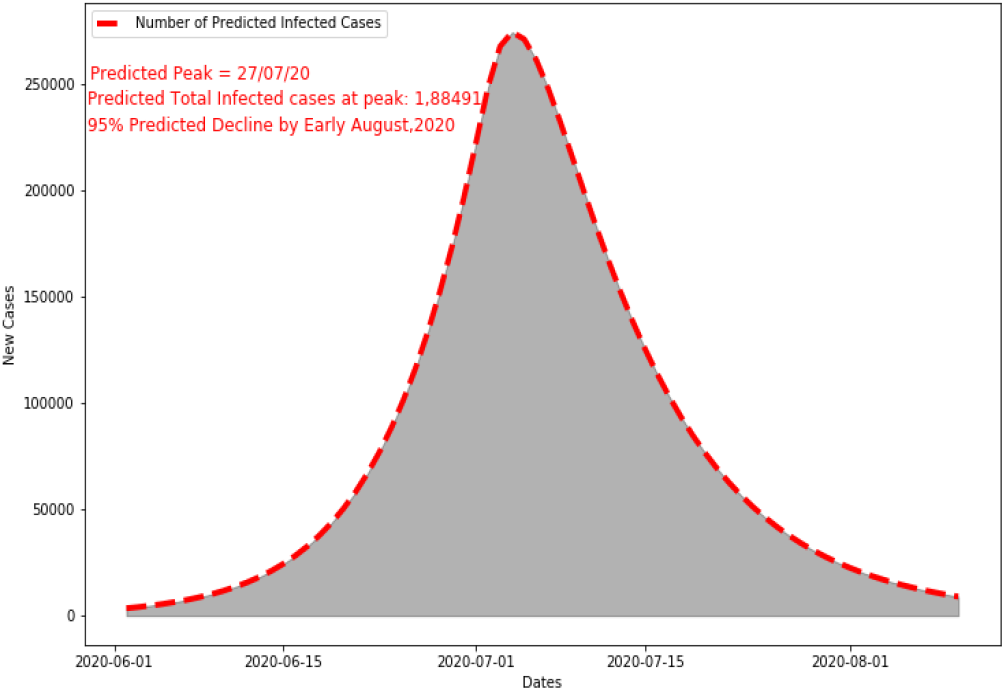
Predicted Peak of New cases In Delhi under Partial to Zero lockdown

## 6 Conclusion

The study done in this work indicated that India is yet to achieve the peak in the spread of Coronavirus disease. The predictive model proposed revealed that it is unlikely to get rid of Covid-19 before end of October 2020 in India. It is ofcourse not good news, but has to be accepted to meet the basic survival. Referring to the model development, it used combination of epidemiological model (SEIR) and mathematical curve fitting method to forecast the impact of the COVID-19 in India. For conventional SEIR to begin simulation, the basic parameters such as, *β*, R_*o*_, *γ, δ*, population are to be fixed. However, in reality these parameters may vary depending on several factors like country, states, lockdown strictness, rules and regulations and social distancing. Keeping them fixed may affect the prediction accuracy of the model. While the factor population of can be easily be identified for the country, the other variable parameters are not easily retrievable. Hence, approximating them through process of “curve fitting” proposed in this work is a good choice. To provide robust learning to the model, the historical data of Coronavirus confirmed cases in India till May 03, 2020 was used. The key reason to choose the data till May 03, 2020 was because till this date India was under strict lockdown phase. Extending the historical data beyond this time for the purpose of learning may have affected the curve parameters. The parameters *β* and R_*o*_ were learnt through the method of “least-square” deployed on SEIR model. This fitting of historical data on SEIR model discovered the possible peak of Corona, i.e., May 14, 2020 in India. However, situation in India was different after May 03, 2020. People in India were given relaxation in movement, migration of labourers, change in diagnostic facilities and many such factors contributed in further progressive spread of the disease. To predict new peak of Coronavirus under this situation is a challenging task since many variable factors influence the Coronavirus infection rate from person to person. Considering this, it was sensible to make the model relearn the situation of spread under partial to zero lockdown phase. This revised learning discovered new peak at July 31, 2020 to be the date of maximum confirmed cases to reach in India. Similar predictions were also done for the capital of India, New Delhi.

The new predictions made for India under partial to zero lockdown are again under assumptions that things will move the way they are currently moving with no variations. However, the predictions for the future may change rapidly with time. Where, different mobility patterns of Indian people, social distancing, Corona testing facility, ban on international travel and group activities will play a key role in increasing or decreasing the infection rate in India. Another factor, which may influence the predictions and affect the distribution of infected cases in future is the virus mutation. However, a study like presented in this work will enable citizens of India, government and medical staff to plan their way forward. In addition to this, contribution of study provides insights on situation of virus spread under full, partial and zero lockdowns. It is the now the awareness of citizens of India to act and behave responsibly to bring life back to normal and mitigate affect of virus from the country.

## Data Availability

Real data set was taken from Kaggle

## References

[1] T. Hiteshi et. al., Coronavirus (COVID-19): ARIMA based time-series analysis to forecast near future. 2004.07859, 2020.

[2] M. Mohsen et. al., Time series modelling to forecast the confirmed and recovered cases of COVID-19. Travel Med. Infect. Dis., no. March, p. 101742, doi: 10.1016/j.tmaid.2020.101742, 2020.

[3] B. Domenico et. al., Application of the ARIMA model on the COVID-2019 epidemic dataset. Data in Brief, 10.1016/j.dib.2020.105340, 2020.

[4] B. Lutfi et. al., Forecasting of COVID-19 Cases and Deaths Using ARIMA Models. MedRxiv, https://doi.org/10.1101/2020.04.17.20069237, 2020.

[5] G. Pandey et. al., SEIR and Regression Model based COVID-19 outbreak predictions in India. 2004.00958, 2020.

[6] P. Narinder et. al., COVID-19 Epidemic Analysis using Machine Learning and Deep Learning Algorithms. MedRxiv, https://doi.org/10.1101/2020.04.08.20057679, 2020.

[7] A. Sina et. al., COVID-19 Outbreak Prediction with Machine Learning. MedRxiv, https://doi.org/10.1101/2020.04.17.20070094, 2020.

[8] Y. Novanto, COVID-19 growth prediction using multivariate long short term memory. 2005.04809, 2020.

[9] T. Shreshth et. al., Predicting the growth and trend of COVID-19 pandemic using machine learning and cloud computing. MedRxiv, https://doi.org/10.1101/2020.05.06.20091900, 2020.

[10] L. Jianxi, Predictive Monitoring of COVID-19. White Paper, May, 2020.

[11] Covid-19 in India, https://www.kaggle.com/sudalairajkumar/covid19-in-india.

[12] Infectious Disease Modelling: Fit Your Model to Coronavirus Data, https://towardsdatascience.com/infectious-disease-modelling-fit-your-model-to-coronavirus-data-2568e672dbc7.

[13] Social Distancing Key to Slowing COVID-19 Spread, https://medium.com/@ngough_bioserendipity/social-distancing-key-to-slowing-covid-19-spread-de3ee86aa34e.

